# Testing, Testing: What SARS-CoV-2 testing services do adults in the United States actually want?

**DOI:** 10.1101/2020.09.15.20195180

**Authors:** Rebecca Zimba, Sarah Kulkarni, Amanda Berry, William You, Chloe Mirzayi, Drew Westmoreland, Angela Parcesepe, Levi Waldron, Madhura Rane, Shivani Kochhar, McKaylee Robertson, Andrew R Maroko, Christian Grov, Denis Nash, for the CHASING COVID Cohort Study Team

## Abstract

**Importance:** Ascertaining preferences for SARS-CoV-2 testing and incorporating findings into the design and implementation of strategies for delivering testing services may enhance testing uptake and engagement, a prerequisite to reducing onward transmission.

**Objective:** To determine important drivers of decisions to obtain a SARS-CoV-2 test in the context of increasing community transmission.

**Design:** A discrete choice experiment (DCE) was used to assess the relative importance of type of SARS-CoV-2 test, specimen type, testing venue, and results turnaround time. Uptake of an optimized testing scenario was simulated relative to the current typical testing scenario of polymerase chain reaction (PCR) via nasopharyngeal (NP) swab in a provider’s office or urgent care clinic with results in >5 days.

**Setting:** Online survey, embedded in an existing cohort study, conducted during July 30 - September 8, 2020.

**Participants:** Participants (n=4,793) were enrolled in the CHASING COVID Cohort Study, a national longitudinal cohort of adults >18 years residing in the 50 US states, Washington, DC, Puerto Rico, or Guam.

**Main Outcome(s) and Measure(s):** Relative importance of SARS-CoV-2 testing method attributes, utilities of specific attribute levels, and probability of choosing a testing scenario based on preferences estimated from the DCE, the current typical testing option, or choosing not to test.

**Results:** Turnaround time for test results had the highest relative importance (30.4%), followed by test type (28.3%), specimen type (26.2%), and venue (15.0%). Participants preferred fast results on both past and current infection and using a noninvasive specimen, preferably collected at home. Simulations suggested that providing immediate or same day test results, providing both PCR and serology, or collecting oral specimens would substantially increase testing uptake over the current typical testing option. Simulated uptake of a hypothetical testing scenario of PCR and serology via a saliva sample at a pharmacy with same day results was 97.7%, compared to 0.6% for the current typical testing scenario, with 1.8% opting for no test.

**Conclusions and Relevance:** Testing strategies that offer both PCR and serology with non-invasive methods and rapid turnaround time would likely have the most uptake and engagement among residents in communities with increasing community transmission of SARS-CoV-2.

## Introduction

The CDC recently estimated that for every case of SARS-CoV-2 infection diagnosed in the U.S., an additional 10 are undiagnosed.^1^ Detecting a higher proportion of people with active infection via widespread testing is a prerequisite to achieving the public health goals of controlling transmission of SARS-CoV-2.^2,3^ However, limited access to and uptake of testing for many in the U.S., combined with lengthy result turnaround time, severely hampers pandemic control efforts, which require timely detection, isolation and quarantine. While recent increases in testing are promising,^4^ some models^5^ suggest a shortfall, and important populations may still be unreached.^6^ Understanding factors that may influence an individual’s decision to seek testing can help enhance and sustain uptake of SARS-CoV-2 testing when, where and among whom it is needed most for public health purposes. These factors include individual preferences for different types of testing services, which have not been systematically ascertained or incorporated into testing service delivery.

## Methods

To identify the most preferred SARS-CoV-2 testing scenarios for individuals, we conducted a discrete choice experiment (DCE)^7,8^ in a U.S. national longitudinal cohort of adults being followed for SARS-CoV-2 seroconversion and other related outcomes. DCEs are a powerful tool to identify the most preferred attributes in populations being targeted for health interventions, and can inform strategies to increase their uptake and engagement.

### Study Population

We invited all participants of the CHASING COVID Cohort (C3) Study^9^ who completed a recent routine follow-up assessment (n=5,098) to participate in the DCE. C3 Study participants were recruited online using internet-based strategies, including via referral and social media advertisements.^9^ Eligibility criteria include being ≥18 years and residing in the US, Puerto Rico, or Guam at enrollment. 4,793 (94% of those invited) completed the DCE July 30-September 8, 2020. A $5 USD Amazon gift card incentive was offered to participants completing the DCE.

### DCE design, analysis, and simulation

The DCE was designed and implemented using Lighthouse Studio 9.8.1 (Sawtooth Software, Provo, UT) and deployed using Sawtooth’s online survey hosting platform. Participants were asked to consider different combinations of SARS-CoV-2 testing service features in a situation where ‘…the number of people hospitalized or dying from coronavirus in your community was increasing.’ Each participant was presented with five choice tasks, each containing two juxtaposed scenarios comprised of different combinations of the testing features (aka attribute levels), and a “None” option if neither testing scenario was appealing or desirable. Testing service attributes included in the DCE are shown in Table 1, and included: type of test, specimen type, testing venue, and results turnaround time (see Supplement eFigure 1 for a sample choice set). The combinations presented and the order of their presentation to each participant were randomized to reduce bias (see eMethods 1 in the Supplement).

**Table 1.** SARS-CoV-2 Testing Discrete Choice Experiment Attributes and Levels.

| Attributes & Levels (abbreviated) | Descriptive Level Text |
| --- | --- |
| **Test** |  |
| Serology | An antibody test that tells you if you've EVER had a COVID-19 infection |
| PCR | A PCR test that tells you if you CURRENTLY have a COVID-19 infection |
| Both tests | BOTH an antibody test (EVER infected) and a PCR test (CURRENTLY infected) |
| **Specimen type** |  |
| Finger prick | A small amount of blood from a finger prick |
| Blood draw | A small tube of blood taken from your arm |
| Cheek | Oral fluid from a swab of the inside of your cheek |
| Spit | A spit sample collected in a small cup |
| Nasal shallow | A SHALLOW swab of the inside of your nostrils |
| NP swab | A DEEP swab that goes far into your nasal passages |
| Urine | A urine sample collected in a small cup |
| **Venue** |  |
| Home collection, receiving & returning kit in mail | You are mailed a package with the test kit, you collect the specimen, and mail it back to the lab |
| Home collection, receiving kit in mail & returning to a collection site | You are mailed a package with the test kit, you collect the specimen, and drop it off at a collection site near your home |
| Doctor's office or urgent care clinic | You go to your doctor's office or an urgent care clinic to have the specimen collected |
| Walk-in community testing site | You go to a walk-in community testing site to have the specimen collected |
| Drive-through community testing site | You go to a drive-thru community testing site to have the specimen collected (you stay in your car) |
| Pharmacy | You go to a local pharmacy to have the specimen collected |
| **Results turnaround time** |  |
| Immediate | Immediately (within 15 minutes) |
| Same day | On the same day |
| 48 hours | Within 48 hours |
| 5 days | Within 5 days |
| > 5 days | > 5 days |
| **None – I wouldn't choose any of these** |  |

*Some combinations of attribute levels were prohibited. For example, a test scenario that included a specimen collected at home and returned to the lab via mail could not also include the immediate test result level.*
*Descriptive text was displayed in the choice exercise.*
Table: CUNY ISPH • Source: Chasing COVID Cohort • Created with Datawrapper

We estimated zero-centered part-worth utilities for each attribute level and overall relative attribute importance using effects coding in a hierarchical-Bayesian model.^10^ We used these estimates to conduct simulations of different strategies against the current typical testing option of a PCR test using a nasopharyngeal (NP) swab in a doctor’s office or urgent care clinic, with results returned in >5 days. We simulated changes in uptake that would result from ‘swapping’ each individual attribute level in Table 1 into the current typical testing option. We also created a testing scenario that optimized preferences across attributes, which included: PCR and serology from a saliva sample collected at a pharmacy with same day results. We then simulated the proportion of participants who would choose this optimized scenario, the current typical testing option, or neither option. Predicted uptake of each testing strategy was simulated using a randomized first choice model,^11,12^ which assumes that each participant would select the scenario that provides them with the highest total utility summed across attributes. DCE data were analyzed and simulations were conducted using Lighthouse Studio 9.8.1.

### Ethical review

The study was approved by the Institutional Review Board at the City University of New York Graduate School of Public Health.

## Results

### Participant demographic characteristics

Participants’ median age was 39 years (IQR 30-53 years). 51.5% identified as female, 45.7% as male, and 2.8% as non-binary. 62.8% identified as Non-Hispanic White, 16.4% as Hispanic, 10.1% as Non-Hispanic Black, 6.9% as Asian, and 3.6% as another race/ethnicity. At enrollment, 29.4% of participants resided in the Northeast, 28.2% in the South, 23.9% in the West, 17.5% in the Midwest, and 0.1% in Puerto Rico or Guam.

### Relative importance of testing service attributes and attribute levels

Results turnaround time had the highest relative importance (30.4%), followed by test type (28.3%), specimen type (26.2%), and venue (15.0%) (see Supplement eTable 1). Participants strongly preferred rapid receipt of results, with progressively weaker preference for slower test results. Within test type, participants showed a strong preference for testing scenarios that detect both current and past infection (see Supplement eTable 1). Participants most preferred testing scenarios that use cheek swab specimens, and least preferred scenarios that require a deep NP swab. There was a preference for at-home self-collection of specimens using kits received and returned via mail; testing in a doctor’s office or urgent care clinic was the least preferred testing venue. Participants chose neither testing option 3.6% of the time.

### Simulation results

Simulating changes in attribute trade-offs individually, providing immediate or same day test results, providing both PCR and serology, or collecting oral specimens would increase testing uptake the most (Figure 1). We also simulated the proportion of participants that would pick the current typical testing scenario versus a scenario with multiple more preferable features: both PCR and serology using a saliva specimen collected at a pharmacy with same day test results. Simulated uptake of this hypothetical scenario was 97.7% compared to 0.6% for the current typical testing scenario, with 1.8% opting for no test when presented with these two choices.

**Figure 1.**
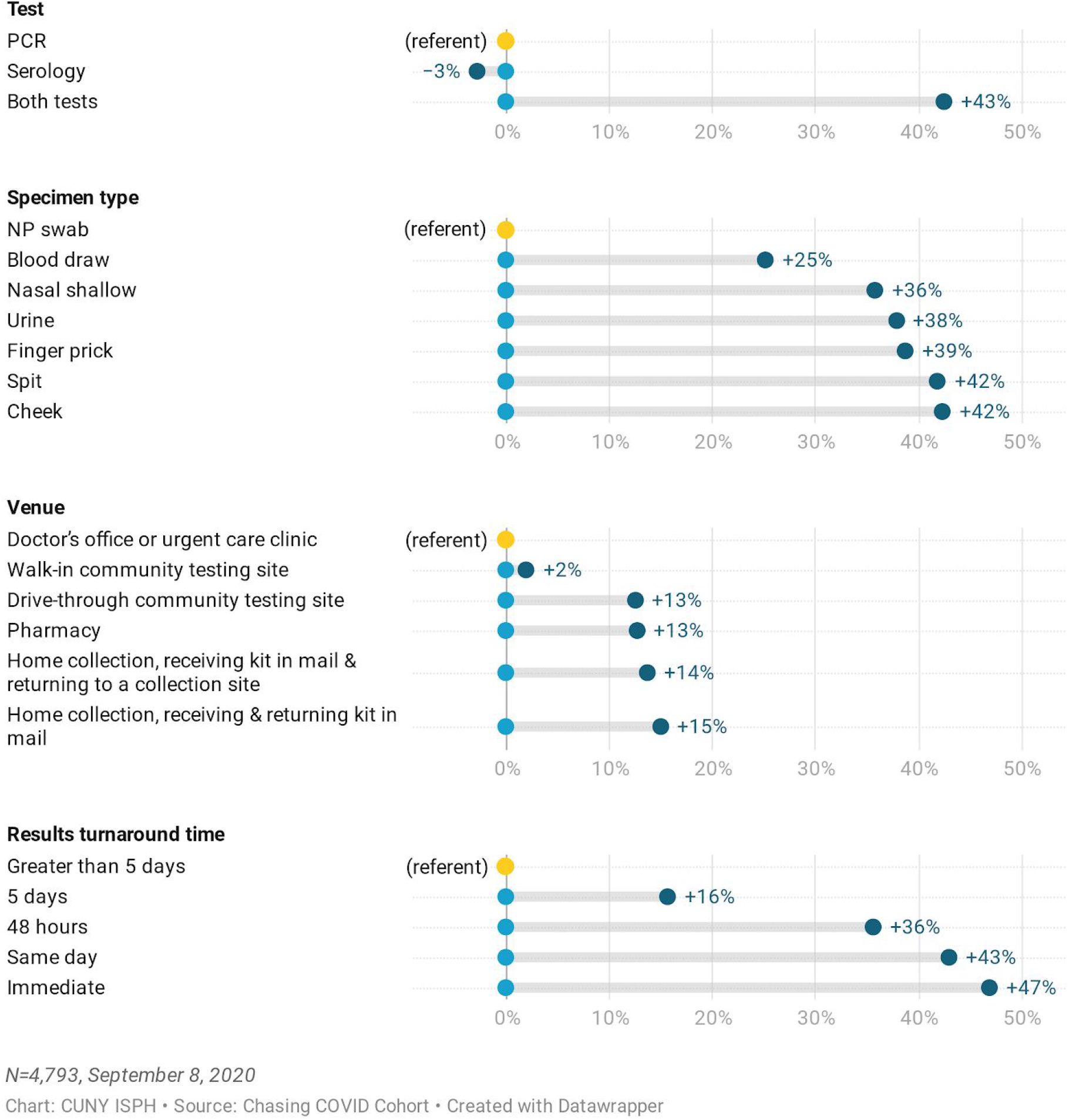
Simulated Changes in SARS-CoV-2 Testing Uptake Relative to the Current Typical Testing Option, by Attribute Level.

## Discussion

These findings suggest that expected advances in SARS-CoV-2 testing technologies, such as rapid, at-home saliva tests, will be highly acceptable and utilized when they become available, particularly in communities with increasing deaths or hospitalizations. Some preferred tests (e.g., at-home rapid antigen tests) may be less sensitive than gold standard diagnostic tests (PCR via NP swab). Nevertheless, these findings are significant from a public health standpoint since its possible that widespread and frequent use of a less sensitive SARS-CoV-2 antigen test could detect much greater numbers of people with active infection—and more quickly—than the current typical testing scenario.^13^ Indeed, our data suggest that NP swabs may be a deterrent to testing, which could be addressed by adding serology or relying on saliva specimens.

Limitations of the study include the omission of other attributes which may influence testing preferences, such as frequency of testing, cost, facility wait times or distance. In addition, the majority of our participants had already completed at-home self-collection of a dried blood spot specimen for our study. Though the venue attribute had the lowest relative importance, this prior experience may have influenced their preferences for venue in the DCE.

To the extent that it is possible to align public health strategies to deliver testing services with the preferences of those being targeted for testing, greater uptake and engagement may be achieved. Additional research is needed to increase SARS-CoV-2 testing uptake in ways that are aligned with the public health goals of the pandemic response, including preferences for engaging in public health interventions following a positive test, such as isolation and contact tracing.^3^

## Data Availability

Requests to access deidentified study data can be made by contacting the investigators directly

## Acknowledgements

We would like to acknowledge the CHASING COVID Cohort Study participants for their contributions to this research.

## eMethods 1

The final DCE design included 500 survey versions in which each level appeared approximately the same number of times as the other levels within each attribute across the five tasks, some level overlap within an attribute was permitted across concepts in the same task, and levels within one attribute were included independently of levels within other attributes—Sawtooth’s Balanced Overlap design.^14,15^ The design was tested with 2500 dummy participants and assuming “None” was chosen in 33% of choice tasks, yielding estimated standard errors ranging from 0.0212 to 0.0489, with an absolute D-efficiency of 1342.39. For comparison, in a completely enumerated design, also with 2500 dummy participants and 33% choosing “None,” the estimated standard errors ranged from 0.0208 to 0.0499 with an absolute D-efficiency of 1380.008; our design’s relative D-efficiency was 97%.

**eFigure 1.**
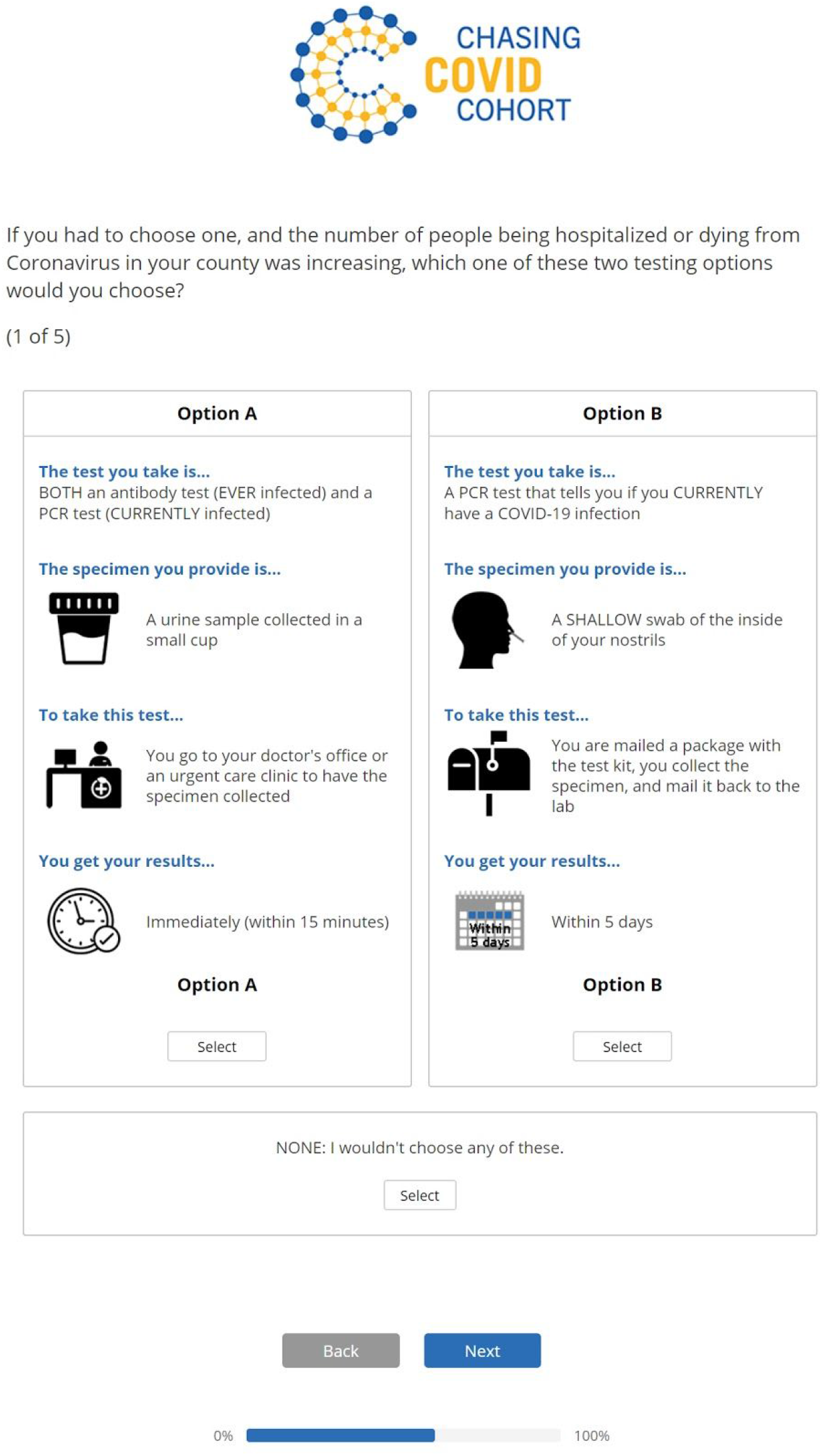
Desktop example of choice task.

**eTable 1.** Part-worth Utilities and Average Relative Attribute Importance for SARS-CoV-2 Testing Features.

| Attributes & Levels | Utility | Lower 95% CI | Upper 95% CI |
| --- | --- | --- | --- |
| **Test** |  |  |  |
| Serology | -37.1 | -37.9 | -36.3 |
| PCR | -24.6 | -25.3 | -24.0 |
| Both tests | 61.7 | 61.0 | 62.4 |
| **Specimen type** |  |  |  |
| Finger prick | 16.3 | 15.7 | 16.8 |
| Blood draw | -19.4 | -20.1 | -18.8 |
| Cheek | 24.9 | 24.5 | 25.3 |
| Spit | 25.1 | 24.5 | 25.6 |
| Nasal shallow | 3.4 | 2.9 | 3.8 |
| NP swab | -63.4 | -64.3 | -62.4 |
| Urine | 13.2 | 12.7 | 13.7 |
| **Venue** |  |  |  |
| Home collection, receiving kit in mail & returning kit in mail | 13.3 | 12.5 | 14.2 |
| Home collection, receiving kit in mail & returning to a collection site | 9.6 | 8.9 | 10.3 |
| Doctor's office or urgent care clinic | -14.4 | -15.0 | -13.8 |
| Walk-in community testing site | -16.8 | -17.4 | -16.2 |
| Drive-through community testing site | 4.8 | 4.3 | 5.3 |
| Pharmacy | 3.5 | 3.1 | 3.9 |
| **Results turnaround time** |  |  |  |
| Immediate | 56.5 | 55.8 | 57.2 |
| Same day | 35.3 | 34.8 | 35.8 |
| 48 hours | 6.7 | 6.4 | 7.0 |
| 5 days | -37.8 | -38.3 | -37.2 |
| GT 5 days | -60.7 | -61.6 | -59.8 |
| **None** | **-228.7** | **-231.0** | **-226.4** |
| Attribute | Relative Importance | Lower 95% CI | Upper 95% CI |
| Test | 28.3% | 28.1% | 28.6% |
| Specimen type | 26.2% | 26.0% | 26.5% |
| Venue | 15.0% | 14.8% | 15.3% |
| Results turnaround time | 30.4% | 30.1% | 30.7% |

N=4,793, September 8, 2020
Table: CUNY ISPH • Source: Chasing COVID Cohort • Created with Datawrapper

